# Balancing Offloading and Comfort: The Effect of Forefoot Rocker Curvature on In-Shoe Plantar Pressure and Gait Kinematics

**DOI:** 10.64898/2026.09.05.26362321

**Authors:** Kamran Shakir, Hadi Sarlak, Anne Sturkenboom, Giulia Rogati, Jaap J. Van Netten, Alberto Leardini, Lisa Berti, Paolo Caravaggi

## Abstract

Therapeutic footwear with rocker soles is widely used to reduce plantar pressure in individuals at risk of diabetic foot ulceration. Of the various rocker sole characteristics, apex position, rocker angle, and apex angle have been extensively studied; however, the effect of forefoot rocker curvature has been seldom investigated. The aim of this study was to assess the isolated effect of forefoot rocker curvature on plantar pressure distribution, lower limb gait kinematics, and comfort.

Ten healthy adults walked in a modular footwear setup with three interchangeable midsoles featuring a straight-line rocker profile (CRi), a rocker radius of 60% (CR60), and 45% (CR45) of the shoe length. In-shoe plantar pressure was measured using 99-sensor capacitive insoles. Kinematics of lower limb joints were measured via inertial measurement units. Comfort-related parameters were assessed using a Visual Analog Scale. Data were analyzed using non-parametric tests.

CRi demonstrated significantly lower pressure at the metatarsal heads than CR60 and CR45 (kPa; median[Q1=Q3]; mean pressure=80 [67–81]; peak pressure=151 [139–169]; p=0.002). CR45 showed significantly lower mean pressure and pressure-time integral in the midfoot than CRi (p=0.002). CR60 showed the largest maximum dorsiflexion angle (deg, median [Q1-Q3]: CR60=17.8[13.4-23.5]; p<0.001) and range of motion (deg: CR60=20.3[18.8-23.3]; p=0.007). CRi was perceived as the least comfortable rocker condition (score 0-100; median[Q1-Q3]; overall comfort=44 [35-57]; p=0.008).

A straight rocker maximizes offloading at the expense of ankle kinematics and comfort; lower rocker radii enhance comfort, thus making the rocker curvature a key design parameter for therapeutic footwear.

## Introduction

Therapeutic footwear plays a vital role in clinical care for individuals with altered foot biomechanics, increased plantar pressure, structural deformities, reduced protective sensation, and a higher risk of plantar tissue damage. Clinical use cases include treating and managing diabetic foot ulcers, rehabilitation, (inactive) Charcot deformity support, and plantar fasciitis (1–4). Since excessive plantar pressure is strongly associated with tissue breakdown and ulcer formation (5,6) design features that redistribute load during gait are central to therapeutic footwear performance.

Rocker soles are one of the most common designs used for offloading in therapeutic footwear (7). By modifying the shoe’s rollover, rocker soles can alter the progression of the center of pressure, reduce loading beneath specific plantar regions, and facilitate forward progression during gait (7). The geometric determinants of forefoot rockers are apex position, rocker angle, apex angle, and rocker radius (Figure 1). An apex position of 50–65% of shoe length, a rocker angle of 15–25 deg, and an apex angle of 90–95 deg have been shown to result in optimal plantar offloading (8–10).

**Figure 1.**
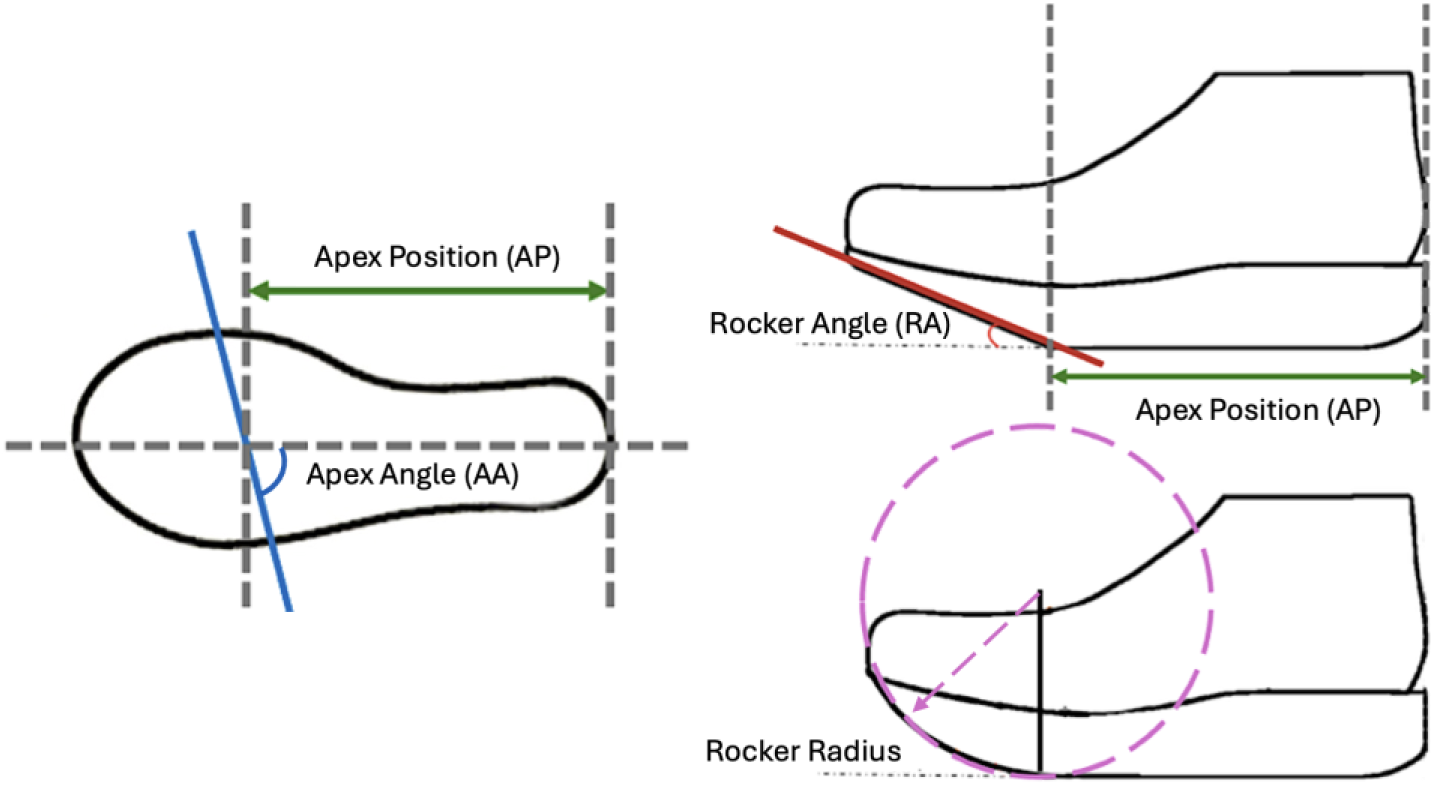
– Schematic representation of the main geometrical parameters used to characterize a forefoot rocker midsole.

However, the effects of the forefoot rocker curvature on biomechanical parameters have been seldom investigated (8,9). Due to its position, the shape of the midsole from the rocker apex to the toe box likely affects the smoothness of plantar load transfer from midstance to toe-off. A smaller rocker radius has been reported to reduce ankle dorsiflexion and plantar-flexion moments in gait (11). Using the same setup, a high-stiffness rocker showed increased peak pressure at the first toe and heel region and decreased at the distal forefoot with respect to a low-stiffness rocker with the same rocker radius. Since rocker curvature may influence both plantar pressure redistribution and perceived comfort, understanding its specific role is important for optimizing therapeutic footwear design aligned with clinical and manufacturing priorities (12).

In general, the effect of the rocker shape on biomechanical parameters has been confounded by the simultaneous change of other design parameters (11) such as the shoe upper design, fit, insole configuration and sensor placement. To address this methodological limitation, a Modular Footwear Setup (MFS) has been recently developed, comprising a flexible shoe upper which can be fitted with interchangeable midsoles (13).

Therefore, the aim of this study was to use the MFS to investigate the effect of forefoot rocker curvature on plantar pressure distribution, lower limb joint kinematics, and perceived comfort in a cohort of healthy participants.

## Methods

### Population

Ten healthy adults (5 M, 5 F; age = 33.2±9.2 yrs; height = 1.73±0.11 m; weight = 65.0±14.1 kg; BMI = 21.5±2.8 kg*m^-2^) were recruited for this study. The sample size was chosen based on testing feasibility and is similar to other biomechanics studies with repeated-measures designs (11,14). Inclusion criteria comprised age 18-70 years, body mass index below 35 kg/m², and compatibility with available shoe sizes (37 and 43 EU). Individuals with prior lower-limb surgery or musculoskeletal deformities were excluded. The study received approval from the local ethics committee (AVEC #647/2025/Sper/IOR). All participants were informed about the data collection process and signed informed consent forms.

### Experimental Setup

A previously developed system for the experimental assessment of footwear interventions, namely the MFS (13), was used in this study (Figure 2A). Three forefoot-rocker midsoles were tested in this study: *CRi*, a straight-line rocker connecting the rocker apex to the toe box (Figure 3, top); *CR60*, a rocker with a radius equal to 60% of the shoe length (Figure 3, middle), and *CR45*, a rocker with a radius equal to 45% of the shoe length (Figure 3, bottom). The midsoles were designed in SolidWorks (version 2025.1, Dassault Systèmes SolidWorks Corp., US) and manufactured using high-density ethylene-vinyl acetate (Asker C 70-75) by trained footwear professionals (Vibram S.p.A, Albizzate, Italy). The three midsoles had the same apex position (65% of the shoe length) and apex angle (90deg). The rearfoot had a rocker (radius = 50 mm) at 10% of the shoe length. The resulting rocker angle of CRi was 25 deg.

**Figure 2.**
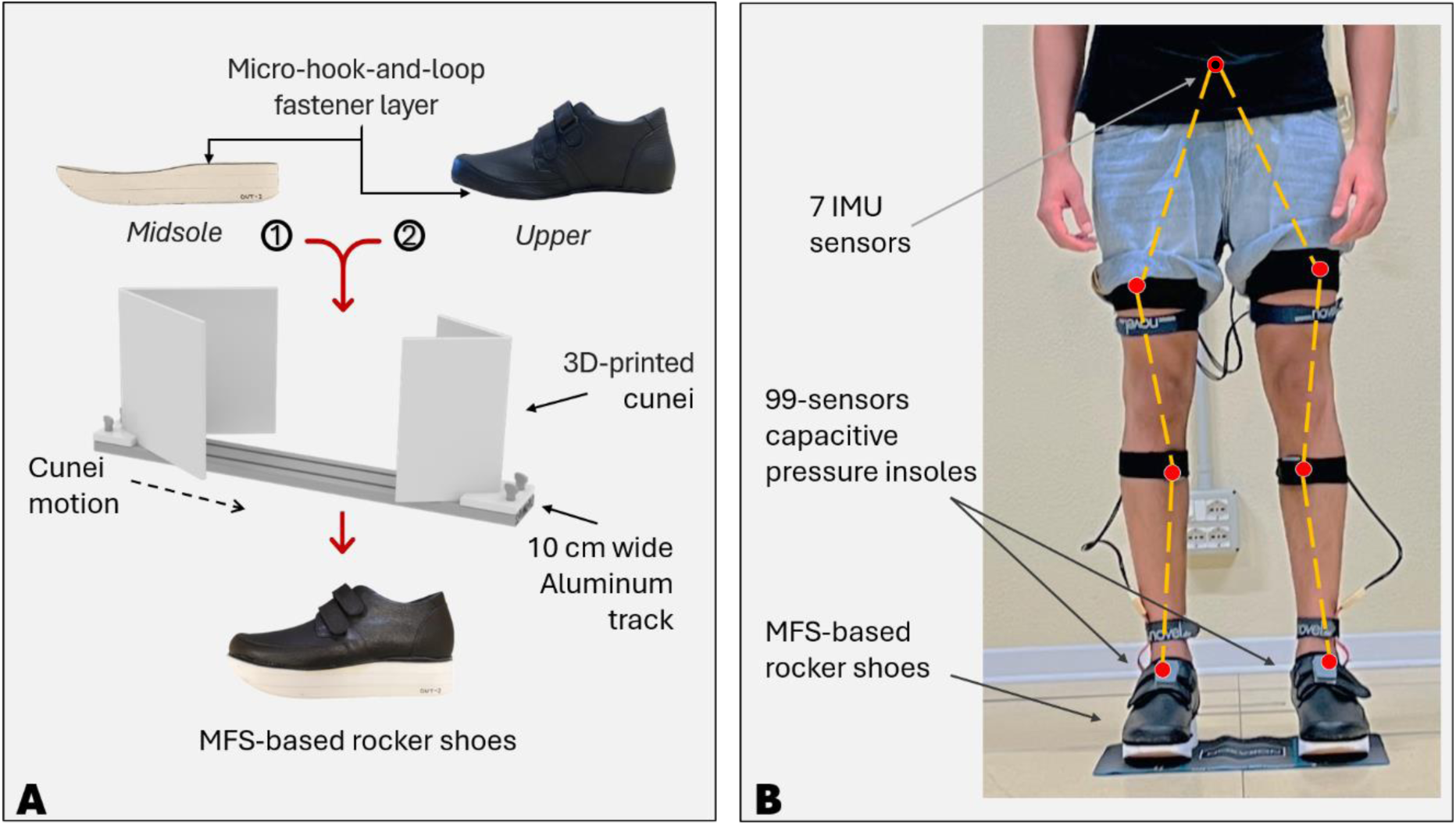
– Diagram of the experimental setup. A: 3D printed cunei allow alignment of the midsole to the upper to change the midsole interventions (13). B: A participant wearing the MFS fitted with a pair of midsoles instrumented with in-shoe pressure insoles and with 7 IMUs in the lower limb (the IMU on the pelvis is attached posteriorly, approximately on S1).

**Figure 3.**
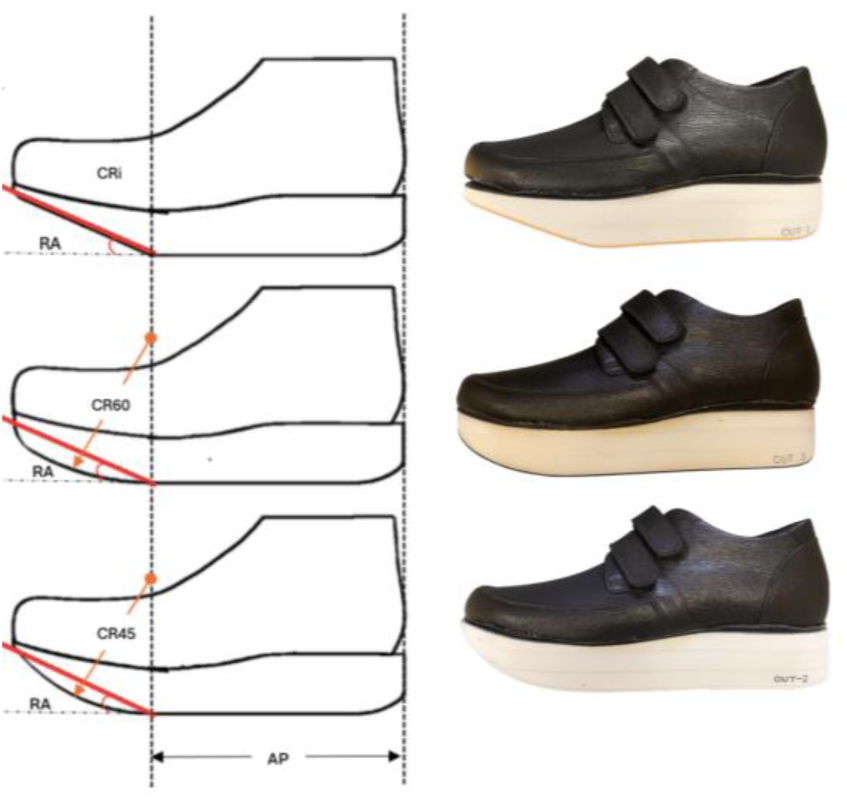
– Schematic representation and lateral photo of the modular footwear setup fitted with the three rocker midsoles. Top: CRi, middle: CR60, and bottom: CR45. The other rocker parameters, apex position and apex angle, were kept constant across all conditions.

Plantar pressure was recorded using two 99-capacitive sensor insoles (Pedar-X, Novel GmbH, Germany) sampling at 100 Hz. Data were analyzed in six regions of interest: whole foot, rearfoot, midfoot, forefoot, metatarsal heads (MTHs), and toes. Mean pressure (MP), peak pressure (PP), and pressure-time integral (PTI) were calculated for each region using a custom MATLAB software (MathWorks, US). Participants were instrumented with 7 Inertial Measurement Units (IMUs, Ultium Motion, Noraxon, US) sampling at 200 Hz. Four IMUs were attached to the shanks and thighs of the left and right legs using Velcro straps. One IMU was attached to the sacrum (approximately on S1), and two were attached to the left and right shoes using double-sided adhesive tape (Figure 2B). Each participant scored the perceived overall comfort, walking stability, and push-off comfort for each shoe condition via a Visual Analog Scale (VAS) questionnaire (Appendix A).

### Experimental Protocol

The participants were instrumented with the IMUs and wore the uppers fitted with the pressure insoles. The selected midsoles were attached to the uppers using the alignment device (13) (Figure 2A). The order of rocker conditions was randomized using the RANDI MATLAB function, and the condition was not revealed to the subject. Participants were given 1 minute to familiarize with each footwear prior to data acquisition. Participants walked at a self-selected comfortable speed along a 10 m laboratory walkway. Walking speed was recorded using a stopwatch. The trial was repeated whenever the speed deviated by over ±10% with respect to that of the initial trial. An average of 11 right-foot steps were analysed for each rocker condition. A VAS questionnaire for comfort was completed after walking in each condition with the opportunity to edit the previous marks.

### Statistics

Non-parametric Friedman tests were used to assess the effect of the rocker condition on biomechanical parameters and comfort. Wilcoxon signed-rank test with Bonferroni correction (α = 0.017) was used for pairwise comparisons. Kendall’s W was used to calculate the effect size and r for pairwise tests. Statistical Parametric Mapping (15) was used to assess differences in temporal profiles of time-normalized joint rotations between the three rocker conditions.

## Results

No difference in walking speed (m*s⁻¹) was observed among the three rocker conditions (median speed [Q1 – Q3]: CRi = 1.19 [1.17 – 1.30], CR60 = 1.21 [1.18 – 1.32], CR45 = 1.22 [1.17 – 1.31]; Friedman’s p = 0.67).

### The effect of rocker curvature on in-shoe plantar pressure

The rockers’ profile significantly affected the pressure parameters in several foot regions (Table 1). In particular, CRi resulted in the greatest offloading at the MTH region, showing significantly lower MP (kPa, median [Q1–Q3]: CRi=80 [67–81]; p=0.001), PP (kPa: CRi=151 [139–169]; p=0.001), and PTI (kPa*s: CRi=60 [48–63]; p=0.001) than the two curved rockers. On the contrary, CRi resulted in larger MP at midfoot (CRi=59 [46–71]; p=0.007) and PTI (KPa*s: CRi=45 [35–55]; p=0.014) and larger PP at rearfoot (kPa: CRi=261 [233–296]; p=0.025). CR45 reduced cumulative midfoot loading (i.e., PTI) relative to CRi, while CR60 resulted in a minimal reduction of PP at the rearfoot, but both increased pressure at the MTHs (see Table 1).

**Table 1.** – Median (IQR) values and statistical test results comparing the three rocker profiles. (*) indicate statistical significance; post-hoc with Bonferroni correction (α = 0.017).

|  |  | Median [Q1–Q3] |  |  | Fried-<br>man’s<br>*p* | Kend-<br>all’s<br>*W* | Wilcoxon pairwise *p*-values and effect size in *r* |  |  |  |  |  |
| --- | --- | --- | --- | --- | --- | --- | --- | --- | --- | --- | --- | --- |
|  |  | CRi | CR60 | CR45 |  |  | CRi vs CR45 |  | CRi vs CR60 |  | CR60 vs CR45 |  |
|  |  |  |  |  |  |  | *p* | *r* | *p* | *r* | *p* | *r* |
| MP<br>[kPa] | Whole<br>foot | 147 [129–<br>177] | 160 [138–<br>178] | 154 [138–<br>171] | 0.122 | 0.21 | 0.193 | 0.29 | 0.020 | 0.52 | 1 | 0.00 |
|  | Rearfoot | 109 [96–<br>125] | 102 [93–<br>115] | 110 [95–<br>115] | **0.045\*** | 0.31 | 0.232 | 0.27 | 0.049 | 0.44 | 0.131 | 0.34 |
|  | Midfoot | 59 [46–<br>71] | 56 [42–<br>67] | 55 [39–<br>64] | **0.007\*** | 0.49 | **0.002\*** | 0.69 | 0.064 | 0.41 | 0.770 | 0.07 |
|  | Forefoot | 85 [79–<br>94] | 97 [86–<br>111] | 95 [87–<br>99] | **0.045\*** | 0.31 | 0.193 | 0.29 | 0.020 | 0.52 | 0.492 | 0.15 |
|  | MTH | 80 [67–81] | 91 [83–102] | 92 [82–94] | **0.001\*** | 0.75 | **0.002\*** | 0.69 | **0.002\*** | 0.69 | 0.557 | 0.13 |
|  | Toes | 67 [56–84] | 72 [61–90] | 73 [65–77] | 0.497 | 0.07 | 0.770 | 0.07 | 0.770 | 0.07 | 0.322 | 0.22 |
| **PP**<br>[kPa] | Whole foot | 265 [233–309] | 256 [230–281] | 267 [233–289] | 0.273 | 0.13 | 0.105 | 0.36 | 0.037 | 0.47 | 0.375 | 0.20 |
|  | Rearfoot | 261 [233–296] | 246 [228–277] | 259 [230–289] | **0.025\*** | 0.37 | 0.232 | 0.27 | **0.014\*** | 0.55 | 0.105 | 0.36 |
|  | Midfoot | 87 [71–110] | 88 [66–106] | 83 [66–104] | 0.150 | 0.19 | 0.037 | 0.47 | 0.375 | 0.20 | 0.625 | 0.11 |
|  | Forefoot | 191 [176–247] | 232 [204–260] | 205 [201–239] | **0.045\*** | 0.31 | 0.432 | 0.18 | 0.193 | 0.29 | 0.084 | 0.39 |
|  | MTH | 151 [139–169] | 209 [182–226] | 202 [170–208] | **0.001\*** | 0.83 | **0.002\*** | 0.69 | **0.002\*** | 0.69 | 0.051 | 0.44 |
|  | Toes | 180 [137–247] | 195 [156–257] | 182 [158–213] | 0.497 | 0.07 | 1.000 | 0.00 | 0.695 | 0.09 | 0.110 | 0.36 |
| **PTI**<br>[kPa*s] | Whole foot | 117 [92–132] | 123 [98–139] | 117 [96–132] | 0.061 | 0.28 | 0.375 | 0.20 | 0.027 | 0.49 | **0.014\*** | 0.55 |
|  | Rearfoot | 83 [69–101] | 78 [68–89] | 80 [67–91] | 0.061 | 0.28 | 0.232 | 0.27 | 0.084 | 0.39 | 0.232 | 0.27 |
|  | Midfoot | 45 [35–55] | 44 [34–51] | 40 [30–49] | **0.014\*** | 0.43 | **0.002\*** | 0.69 | 0.064 | 0.41 | 0.625 | 0.11 |
|  | Forefoot | 68 [55–74] | 71 [63–87] | 70 [61–77] | 0.202 | 0.16 | 0.275 | 0.24 | 0.027 | 0.49 | 0.131 | 0.34 |
|  | MTH | 60 [48–63] | 69 [60–82] | 68 [59–73] | **0.001\*** | 0.76 | **0.002\*** | 0.69 | **0.002\*** | 0.69 | 0.160 | 0.31 |
|  | Toes | 54 [43–62] | 57 [46–67] | 54 [50–58] | 0.741 | 0.03 | 0.770 | 0.07 | 0.695 | 0.09 | 0.232 | 0.27 |

### The effect of rocker curvature on joint kinematics

The rocker profiles significantly affected sagittal-plane ankle joint rotations at late stance (Figure 4). CR60 showed the largest maximum dorsiflexion angle (deg, median [Q1 – Q3]: CRi=12.4 [10.8-16.1]; CR60=17.8 [13.4-23.5]; CR45=15.3 [11.7-21.0]; p<0.001) and range of motion (deg, CRi=18.4 [16.1-20.7]; CR60=20.3 [18.8-23.3]; CR45=21.1 [18.5-23.6]; p=0.007). Statistical parametric mapping analysis of the sagittal-plane ankle joint rotations is reported in Appendix B. No statistically significant differences in sagittal-plane joint rotations were detected between rockers at the hip and knee over the gait cycle (Figure 4).

**Figure 4.**
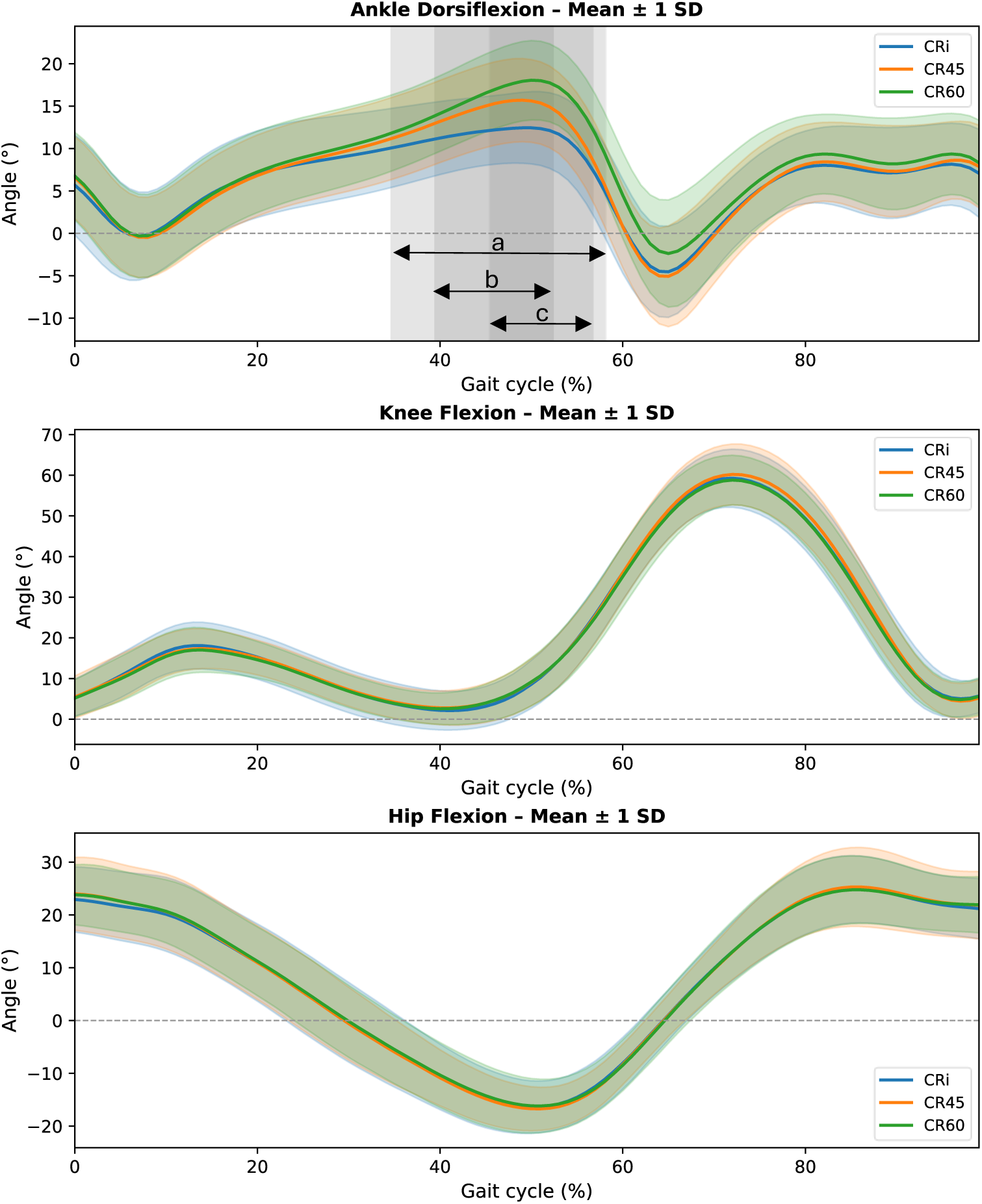
– Ankle (top), knee (middle), and hip (bottom) joint rotations in the sagittal plane (mean ± SD across 10 participants) while walking in each midsole. The shaded patches in ankle (top) joint rotations indicate statistical significance at that specific part of the gait cycle. (a) – CRi vs CR45; (b) – CRi vs CR60; (c) – CR60 vs CR45

### The effect of rocker curvature on perceived comfort

CR60 was perceived as the most comfortable in terms of walking stability and push-off comfort, showing statistically significantly higher scores than CRi (Table 2). CR45 was perceived as significantly more comfortable than CRi.

**Table 2.** – Results of the statistical analyses on Visual Analog Scale (range of 0-100, 100 being the highest) scores of comfort ratings. (*) indicate statistical significance; post-hoc with Bonferroni correction (α = 0.017).

|  | Median [Q1–Q3] |  |  | Friedman’s<br>*p* | Kendall’s<br>*W* | Wilcoxon pairwise *p*-values + effect size in *r* |  |  |  |  |  |
| --- | --- | --- | --- | --- | --- | --- | --- | --- | --- | --- | --- |
|  | CRi | CR60 | CR45 |  |  | CRi vs CR45 |  | CRi vs CR60 |  | CR60 vs CR45 |  |
|  |  |  |  |  |  | *p* | *r* | *p* | *r* | *p* | *r* |
| Overall Comfort | 44<br>[35-57] | 71<br>[56-76] | 64<br>[57-75] | **0.004\*** | 0.562 | **0.008\*** | 0.844 | 0.022 | 0.725 | 0.646 | 0.145 |
| Walking Stability | 38<br>[22-50] | 67<br>[58-79] | 61<br>[46-74] | **0.014\*** | 0.430 | 0.047 | 0.629 | **0.007\*** | 0.854 | 0.241 | 0.371 |
| Push-off Comfort | 27<br>[19-49] | 63<br>[58-70] | 62<br>[48-73] | **0.007\*** | 0.490 | 0.028 | 0.693 | **0.009\*** | 0.822 | 0.508 | 0.210 |

## Discussion

This study explored how forefoot rocker curvature affects biomechanical parameters and comfort in healthy participants. The lowest pressure parameters at the forefoot were observed for the straight-line rocker. This was supported by a large effect size (r=0.69), indicating a consistent offloading effect across participants. CRi showed a proximal shift in loading, evident in higher midfoot MP and PTI than those in CR45 and higher rearfoot PP than in CR60. These results support the albeit-limited evidence from previous investigations that forefoot rocker curvature alone can significantly alter plantar pressure distribution (8,16). Although not statistically significant, a trend for reduced PP at the MTH region was also observed for the smaller rocker radius (PP: CR45<CR60; p=0.051), which appears to be consistent with the association between rocker radii and plantar pressure parameters reported by Ten Wolde et al. (11). This association between geometric and in-shoe pressure parameters is relevant to the prevention of foot complications in people with diabetes, as excessive cumulative plantar stress (associated with PTI) can lead to tissue breakdown and, eventually, to ulceration (17).

The midsole rocker profile significantly affected ankle joint kinematics in gait. CRi altered ankle kinematics in the sagittal plane at late stance, just prior to push-off, by reducing ankle dorsiflexion. This rocker likely induces early plantarflexion, thus initiating the push-off phase earlier than the other conditions. The association between smaller rocker radii and lower ankle dorsiflexion appears to be consistent with what was reported by van Kouwenhove et al. (14).

A trade-off between biomechanical efficacy and perceived comfort was observed with varying rocker curvature, and the two curved rocker profiles were perceived very differently from the straight-line rocker. Although CRi achieved the greatest MTH offloading, it was perceived as the least comfortable. In contrast, CR60 received the highest scores across the three domains. This variation, consistent with earlier reports (10), suggests that the most effective pressure-reducing designs may not be the most acceptable or comfortable option. Since comfort can influence adherence to therapeutic footwear (18,19), a rocker design that balances pressure reduction with walking comfort may offer greater real-world effectiveness than an extreme offloading configuration, due to improved adherence. Comfort outcomes also suggest that smoother rollover profiles may improve user experience. CR45 and CR60 did not differ significantly in comfort despite differences in midfoot loading, suggesting that once rollover becomes sufficiently smooth, further changes may offer limited benefits, at least in healthy individuals. This supports the need for region-specific and patient-specific optimization rather than selecting rocker geometry based solely on maximal forefoot offloading.

To the best of the authors’ knowledge, this is the first study to simultaneously analyze the effects of rocker curvature on lower limb joint kinematics, in-shoe plantar pressure, and comfort. The investigation was supported by a recently developed in-house setup (13) designed to improve measurement repeatability and to minimize the effect of confounding factors such as fit variations, upper construction, and sensor placement. Testing two shoe sizes (EU 37 and 43) further strengthens the robustness of the findings. The results of this study should be assessed considering some limitations. While an effort was made to change only the curvature of the rocker without modifying the other rocker parameters, the nominal rocker angle of 25 deg could be guaranteed only for the straight-line rocker. However, in this investigation, the comfort provided by a smooth curvature (center of curvature right above the rocker apex) and the preservation of the midsole thickness and weight were preferred. In addition, limited sample size, inclusion of healthy participants only, and the limited familiarization period with each footwear condition should be taken into account when interpreting the results of this study. Future research should use larger samples and include clinical populations, such as people with diabetes and peripheral neuropathy, for which plantar offloading is particularly critical.

## Conclusion

According to the present investigation, forefoot rocker curvature significantly affects plantar pressure distribution, ankle dorsiflexion, and perceived comfort. A straight-line rocker maximizes MTH offloading, whereas a moderate curvature radius enhances comfort. These findings suggest that rocker curvature should be considered as a key design parameter to optimize therapeutic footwear in terms of biomechanical effectiveness, user acceptability, and adherence.

## Acknowledgements

The authors would like to thank Filippo Goi (Vibram, S.p.A, Italy) and Giorgia Sartorato (Podartis SRL, Italy) for their help with producing the MFS parts used in this study. This project has received funding from the European Union’s Horizon 2020 research and innovation program under the Marie Skłodowska-Curie grant agreement no. 101073533 (DIALECT: Diabetes Lower Extremity Complications Research and Training Network in Foot Ulcer and Amputation Prevention).

## Disclosure

### Ethics approval and consent to participate

This study received approval from the local ethical committee (AVEC #647/2025/Sper/IOR). All participants were informed about the data collection process and provided informed consent to participate.

### Consent for publication

Not applicable

### Availability of data and materials

Data produced during this study could be made available upon reasonable request from the corresponding author.

### Authors’ contributions

K.S. and H.S. contributed to the conceptualization of the study, data curation, formal analysis, methodology, validation, and visualization, and were involved in writing the original draft and reviewing and editing the manuscript. A.S. contributed to the data curation, methodology, validation, and was involved in reviewing and editing the manuscript. G.R. helped with the methodology, provided supervision, validation, and review and editing. J.N. contributed to the project administration, supervision, review, and editing. A.L. handled project administration, resources, supervision, validation, review, and editing. L.B. contributed to the project administration, supervision, review, and editing. P.C. was involved in conceptualization, formal analysis, methodology, supervision, validation, and the preparation, review, and editing of the original draft.

## Appendix A

### Questionnaire A1

#### Visual Analogue Scale (VAS) questionnaire for comfort

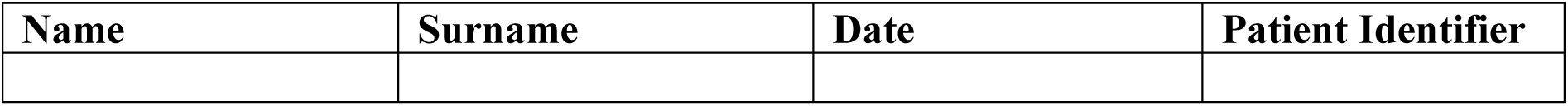

Please indicate your perception of each of the following aspects by marking a vertical line on the horizontal scale below.

– The left end of the line (0) means “not at all / extremely uncomfortable”.
– The right end of the line (100) means “extremely / comfortable”.
– Place your mark anywhere along the line that best represents your perception.

<u>Comfort:</u> how comfortable did you find the shoe during walking?

<u>Stability:</u> how stable did you find the shoe during walking? With stability we mean the extent to which you had the feeling you were able to walk in a stable manner.

<u>Push-off characteristics</u>: how comfortable did the shoe feel during the push-off phase of walking? With push-off we mean the phase where you use your toes to push yourself forward to the next step.

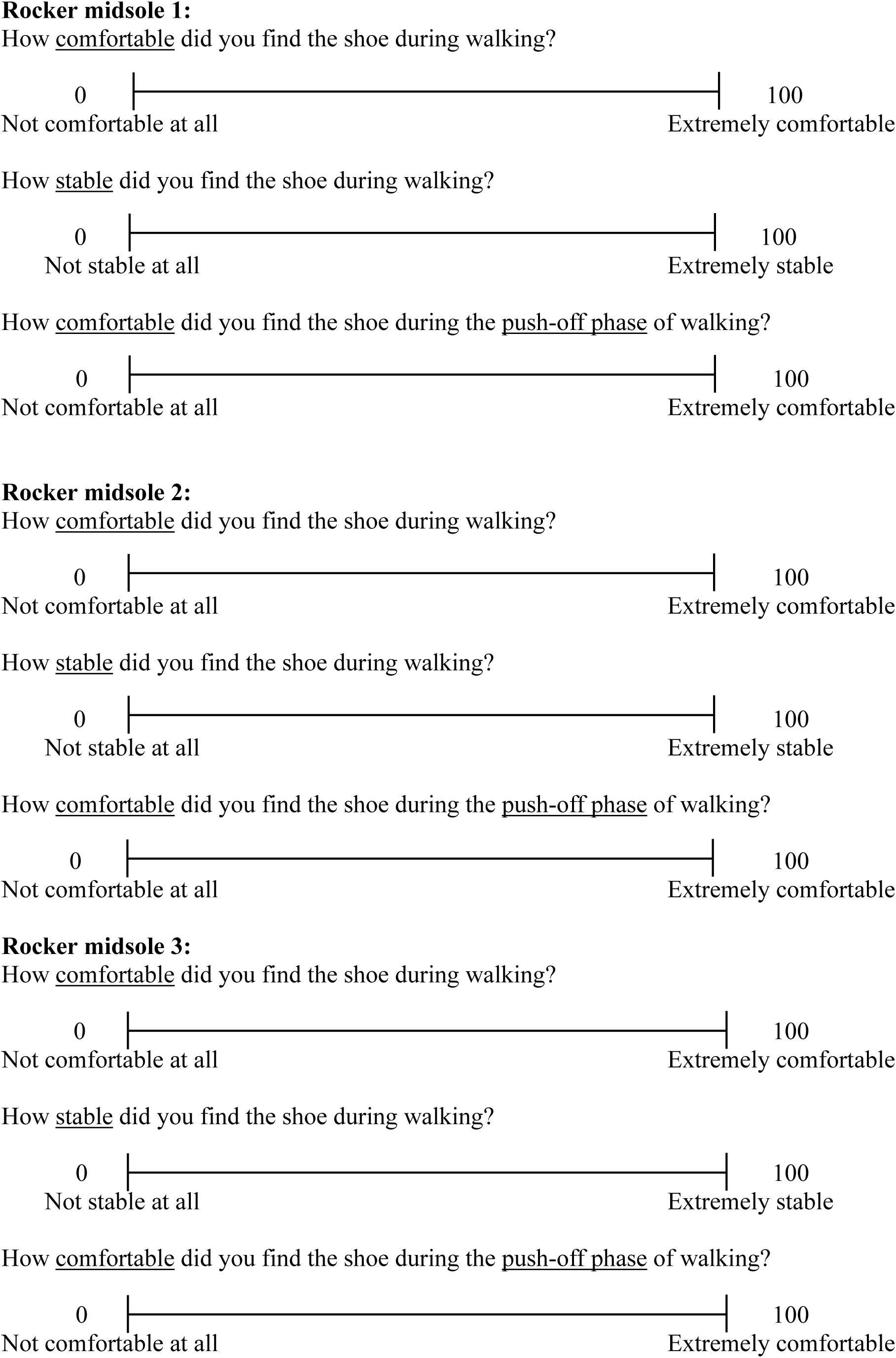

## Appendix B

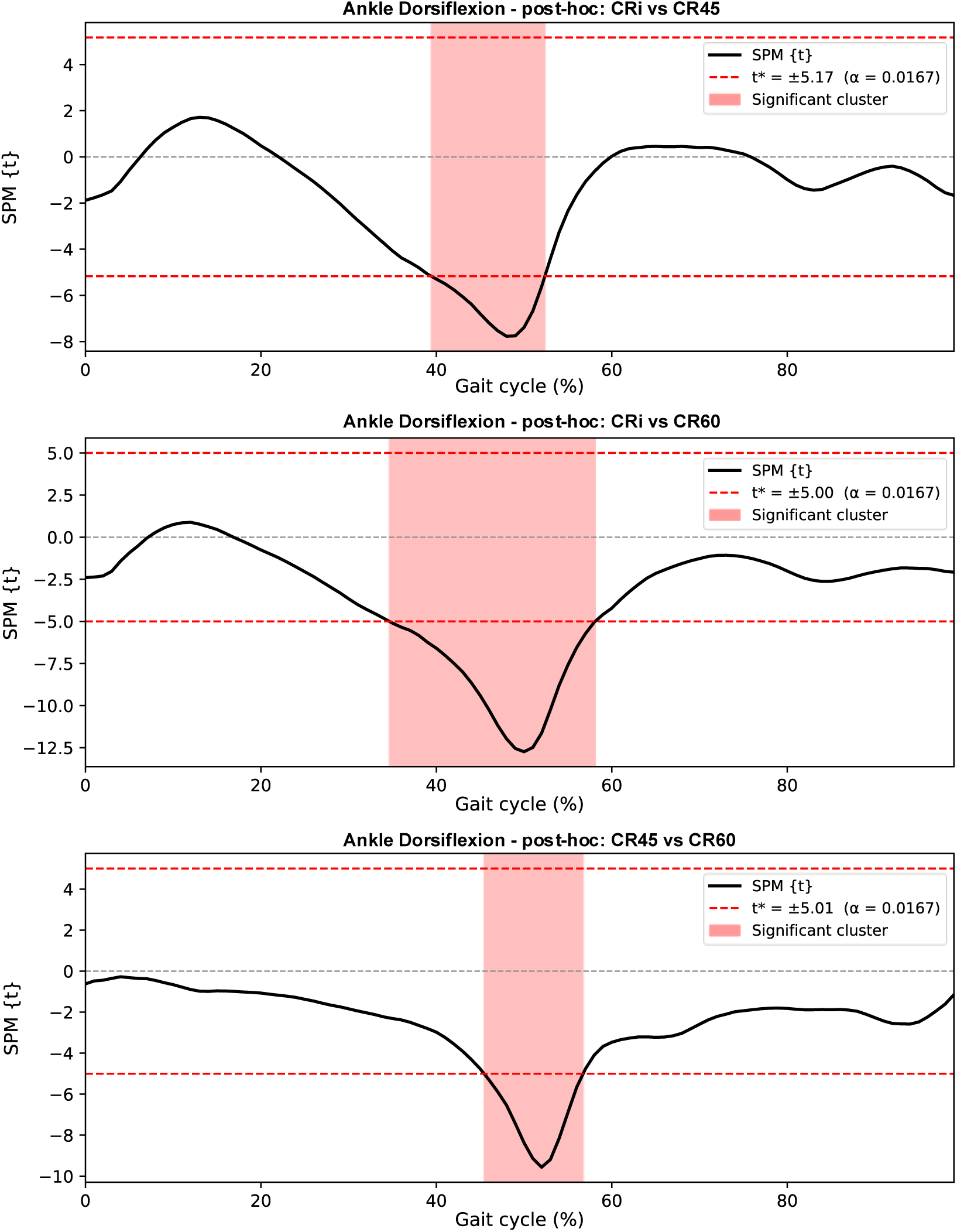

